# Cancer services during the COVID-19 pandemic: systematic review of patients’ and caregivers’ experiences

**DOI:** 10.1101/2021.03.19.21253949

**Authors:** Symran Dhada, Derek Stewart, Ejaz Cheema, Muhammed Abdul Hadi, Vibhu Paudyal

## Abstract

**Background:** Cancer patients have faced intersecting crises in the face of COVID-19 pandemic. This review aimed to examine patients’ and caregivers’ experiences of accessing cancer services during the COVID-19 pandemic and perceived impact of the pandemic on their psychological wellbeing.

**Methods:** A protocol-led (CRD42020214906) systematic review was conducted by searching six databases including EMBASE, MEDLINE and CINAHL for articles published in English-language between 1/2020-12/2020. Data were extracted using a pilot-tested, structured data extraction form. Thematic synthesis of data was undertaken and reported as per the PRISMA guideline.

**Results:** A total of 1110 articles were screened of which 19 studies met the inclusion criteria. Studies originated from 10 different countries including the US, UK, India and China. Several themes were identified which were categorised into seven categories. Postponement and delays in cancer screening and treatment, drug shortages and inadequate nursing care were commonly experienced by patients. Hospital closures, resource constraints, national lockdowns and patient reluctance to use health services because of infection worries contributed to the delay. Financial and social distress, isolation; and spiritual distress due to the uncertainty of rites as well as fulfilment of last wishes were also commonly reported. Caregivers felt anxious about infecting cancer patients with COVID-19.

**Conclusions:** Patients and caregivers experienced extensive impact of COVID-19 on cancer screening, treatment and care, and their own psychological wellbeing. Patient and caregiver views and preferences should be incorporated in ensuring resilient cancer services that can minimise the impact of ongoing and future pandemic on cancer care and mitigate patient fears.

**Protocol Registration:** Published protocol registered with Centre for Review and Dissemination CRD42020214906 (https://www.crd.york.ac.uk/prospero/display_record.php?RecordID=214906)

## Introduction

Coronavirus disease 2019 (COVID-19), caused by the novel severe acute respiratory syndrome coronavirus 2 (SARS-CoV-2) [1], has challenged resilience of healthcare systems and economies globally. The pandemic has placed an immense strain on cancer services, resulting in disruptions to oncology practices contributed both by measures to minimise patient exposure to the virus and strains on health services resources. Such disruptions in services inflicts dual disadvantage to cancer patients as they have poor prognosis compared to non-cancer patients [2]. Overall case fatality rate due to COVID-19 for cancer patients is reported to be 23.4% (95% Cl= 9.7% to 40.5%) compared to 5.9% (95% Cl= 1.9% to 11.7) for non-cancer patients [3]. Data estimates suggest up to 85% reduction in cancer screening services in the US at the height of the first pandemic (March-July 2020) [4]. In the UK, there were 290,000 fewer urgent referrals to cancer services during the first national lockdown (between March and June) [5]. Subsequently, the number of avoidable cancer related deaths are expected to increase substantially6 as early diagnosis and timely access to appropriate treatment modality is critical in ensuring good patient outcomes [2,7,8]. Such interruptions in services can also negatively affect cancer patients undergoing or awaiting a palliative care.

The rapid resurgence of COVID-19 pandemic and diversification of health services to mitigate the impact of the pandemic has led many to question whether cancer has become the ‘forgotten C’[9,10] Cancer services have the difficult task of minimising patient exposure to the virus whilst not compromising cancer outcomes. For patients and caregivers, anxieties and fear may be heightened from psychological perspectives. Whilst current research places an emphasis on linking COVID-19 to treatment adjustments and survival outcomes [3,6-8,11,12], it is important to consider patient and caregiver views and experiences in developing resilient cancer service models for ongoing and future pandemic. This review aimed to examine patients’ and caregivers’ experiences of accessing cancer services during the COVID-19 pandemic and perceived impact of the pandemic on their psychological wellbeing.

## Methods

A systematic review was conducted and reported in accordance with the recommendation of Preferred Reporting Items for Systematic Reviews and Meta-analyses (PRISMA) guidelines [13] based on a published protocol (CRD42020214906). CINAHL, EMBASE, MEDLINE, Google Scholar; and Wellcome Open Research and Authorea (for unpublished studies ongoing peer reviews) databases were searched systematically using medical subject headings (MeSH) and free-text keywords including ‘cancer’, ‘oncology, and ‘Coronavirus’, ‘COVID-19’ and related terms (electronic supplementary material 1) from December 2019 till December 2020. Boolean operators were used to refine the search.

Studies published in English language that explored the views, experiences and perceived impact of COVID-19 on cancer screening and care from the perspective patients/public and their caregivers (both formal and informal) were included. Search was not restricted to any cancer types or participant demography or age. Experiences of cancer screening and care in any setting including cancer related palliative care were included.

Initial screening of titles was conducted independently by two investigators (SD and VP) in order to identify potentially relevant papers. Disagreements were resolved through discussion and a third investigator was consulted where necessary to resolve any disagreements. This was then followed by abstract and full-text screening against the inclusion and exclusion criteria. Data was extracted by one author (SD) and checked by another author (VP) for accuracy using a pilot tested, structured data collection sheet to extract relevant data in relation to views, experiences and perceived impact of COVID-19 on cancer screening, care and psychological impact on patients and carers. Thematic synthesis was adopted to analyse the extracted data and report into categories and themes [14]. A Critical Appraisal Skills Programme (CASP) tool was used to assess the quality of the qualitative studies [15] and the ‘NIH Quality of Observational, Cohort and Cross-Sectional Studies Assessment Tool’ was used to assess the quality of studies using survey methodology [16] Reporting was done as per PRISMA checklist[13] (electronic supplementary material 2).

## Results

### Characteristics and quality of included studies

The initial search yielded 1110 results. A total of nineteen studies [17-35] were included in the final review (figure 1). Studies represented experiences from 10 different countries and were largely based in Italy (n=4) followed by the US (n=3) and India (n=3). Two studies were conducted in the UK and the Netherlands. Five studies used qualitative methods (table 1). The majority of studies employed a survey methodology using questionnaire as data collection tools (n=14) (table 2). Results of quality assessment are presented in electronic supplement 3 and 4.

**Table 1:** Characteristics of qualitative studies and key results.

| Study ID & (Country) | Study aim(s) | Study setting | Study design & methodology | Participant numbers & selection | Key findings |
| --- | --- | --- | --- | --- | --- |
| Dhavale 2020 (India) (22) | To describe the challenges faced by patients and caregivers during the lockdown due to the COVID-19 pandemic. | Palliative care centre. | A qualitative study using an exploratory approach to review case notes followed by framework analysis. | Out of 30 patients, 9 families that had received care services during the lockdown period were invited to participate. | A range of challenges were faced by patients including: physical distress due to lack of availability of medicines and nursing care; emotional distress due to the interruption of cancer treatment; financial and social distress about loss of incomes, isolation; and spiritual distress due to the uncertainty of rites as well as fulfilment of last wishes. Three key themes were identified: swinging on the path of fear to adaptation, left-alone at emotional distances and care system confusion, and decreased quality of care. |
| Mirlashari 2020 (Iran) (29) | To investigate the perspectives of children with cancer and their family in the era of the COVID-19 pandemic. | Paediatric hospital. | Semi-structured, telephone interviews followed by thematic analysis. | 21 participants: five children, thirteen mothers, a father and three paediatric oncology nurses were selected using the purposive sampling technique. | The children expressed their concerns about being exposed to an unknown and enormous threat. They pointed out other significant concerns, such as changes in the treatment process, the lack of effective treatment, and how the disease has become so widespread. |
| Hyland 2020 (United States) (28) | To characterise the behavioural and psychosocial responses of people with advanced lung cancer to the COVID-19 pandemic. | Cancer centre. | Baseline questionnaire and semi-structured interview were conducted and repeated after one month. | 15 patients. | Several themes emerged from the data: cancer as the primary health threat, changes in oncology practice and access to cancer care, awareness of mortality and perceptions of risk, behavioural and psychosocial responses to COVID-19, sense of loss/mourning and positive reinterpretation/greater appreciation of life. All participants reported changing their behaviour in response to COVID-19. |
| AlWaheidi 2020 (Gaza) (18) | To assess whether COVID-19 could lead to further inequity in cancer care and poorer outcomes for Palestinians with cancer. | Tertiary care setting. | Semi-structured qualitative interviews were conducted prior to the introduction of COVID-19. The women were then followed-up in order to examine the changes in health and experiences of care. | 20 women with a breast cancer diagnosis between 2017 and 2018. | A number of women expressed concerns about catching COVID-19 in the hospital setting. New concerns emerged about the impact of COVID-19 on treatment. |
| Casanova 2020 (Italy) (19) | To assess the perception of risks and level of stress concerning COVID-19 amongst young patients with cancer. | Paediatric oncology unit. | Semi-structured qualitative, online questionnaire. | 75 patients: 25 were receiving treatment, 25 were in follow-up and had completed their treatment and 25 were healthy peers. | Whilst the majority of healthy peers did not expect to be affected by the virus, a large proportion of patients (more than those in follow-up), were worried and felt at risk of severe complications. Most responders in all three groups reported that they changed their daily habits. |

**Table 2:**
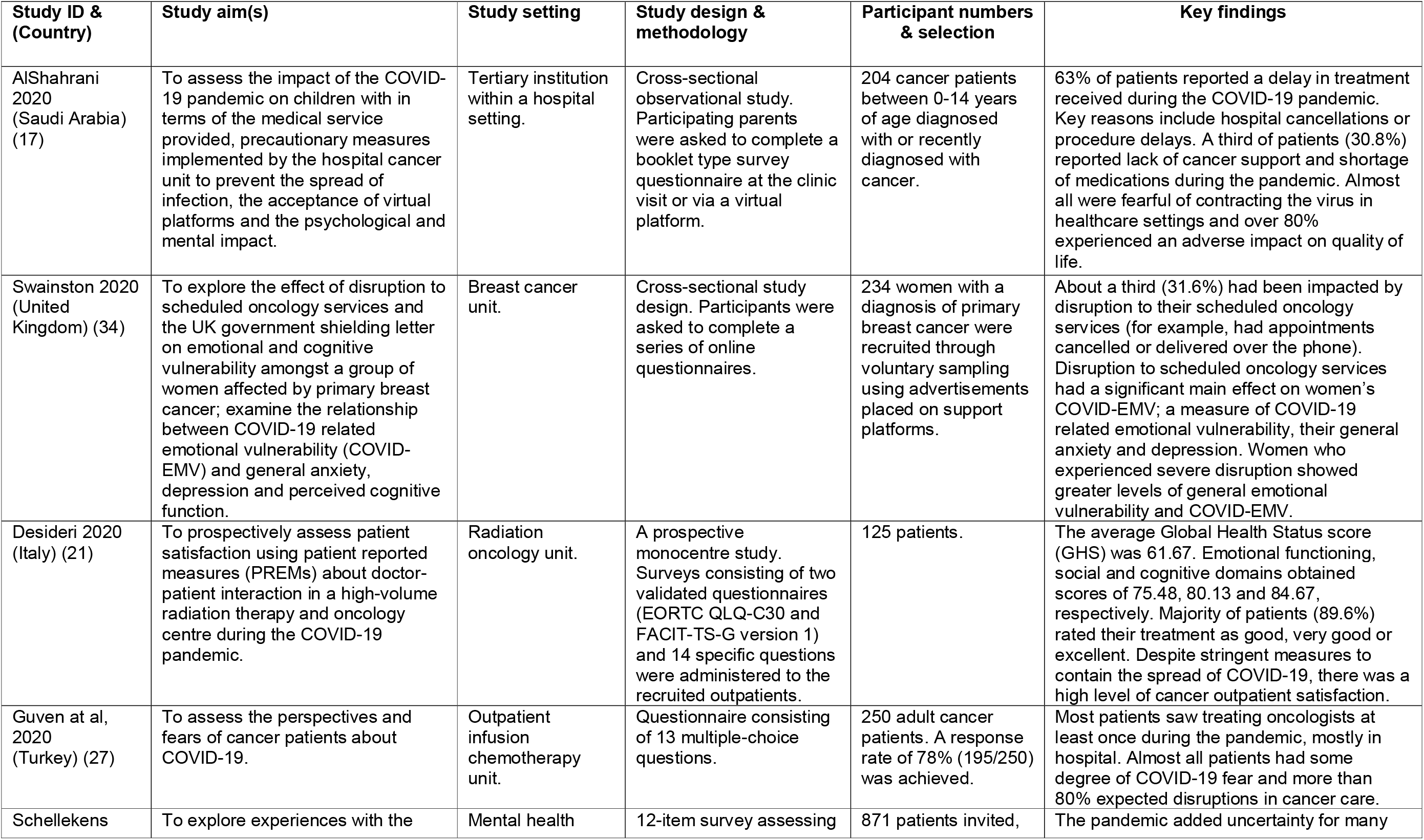
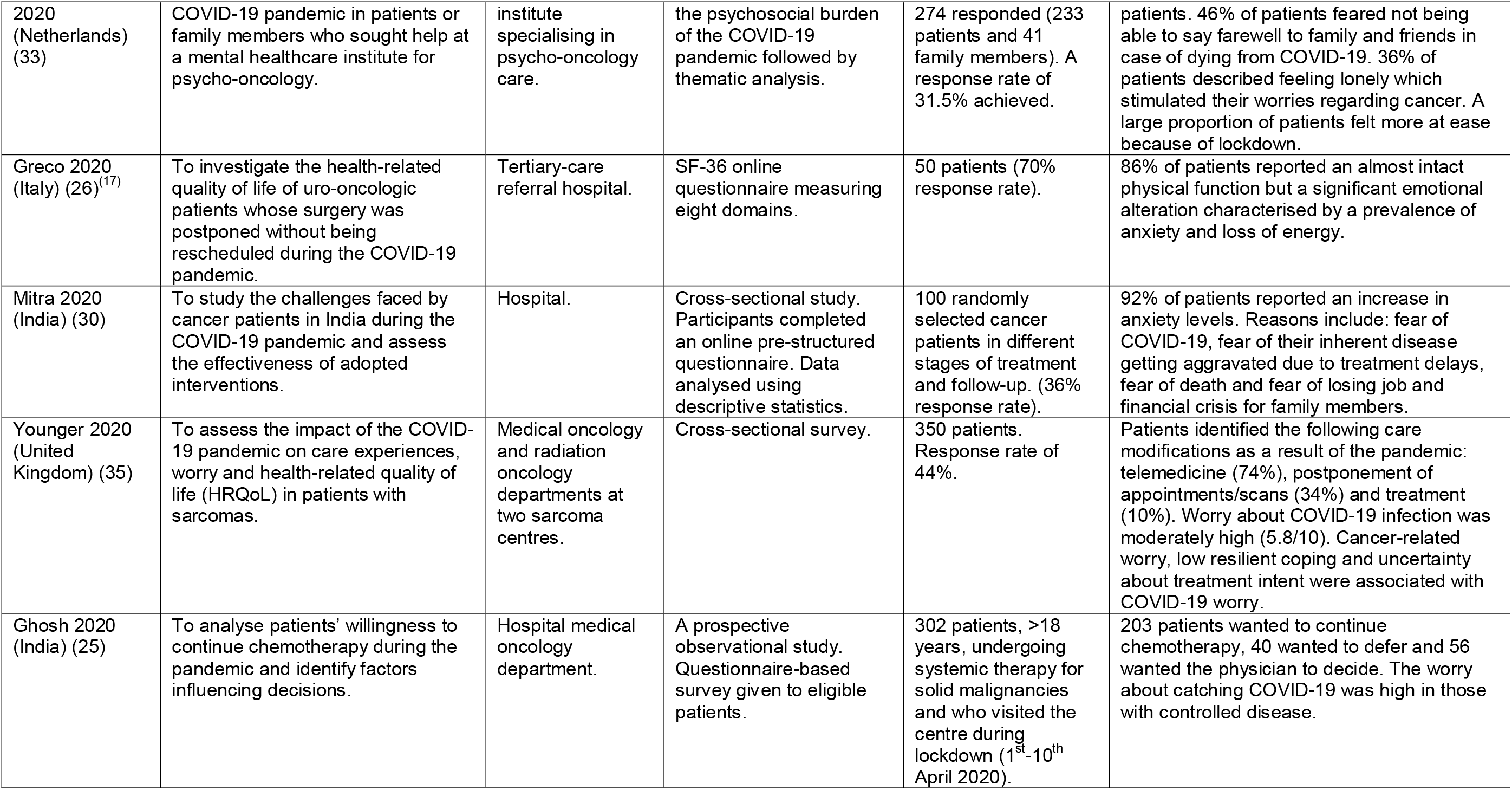
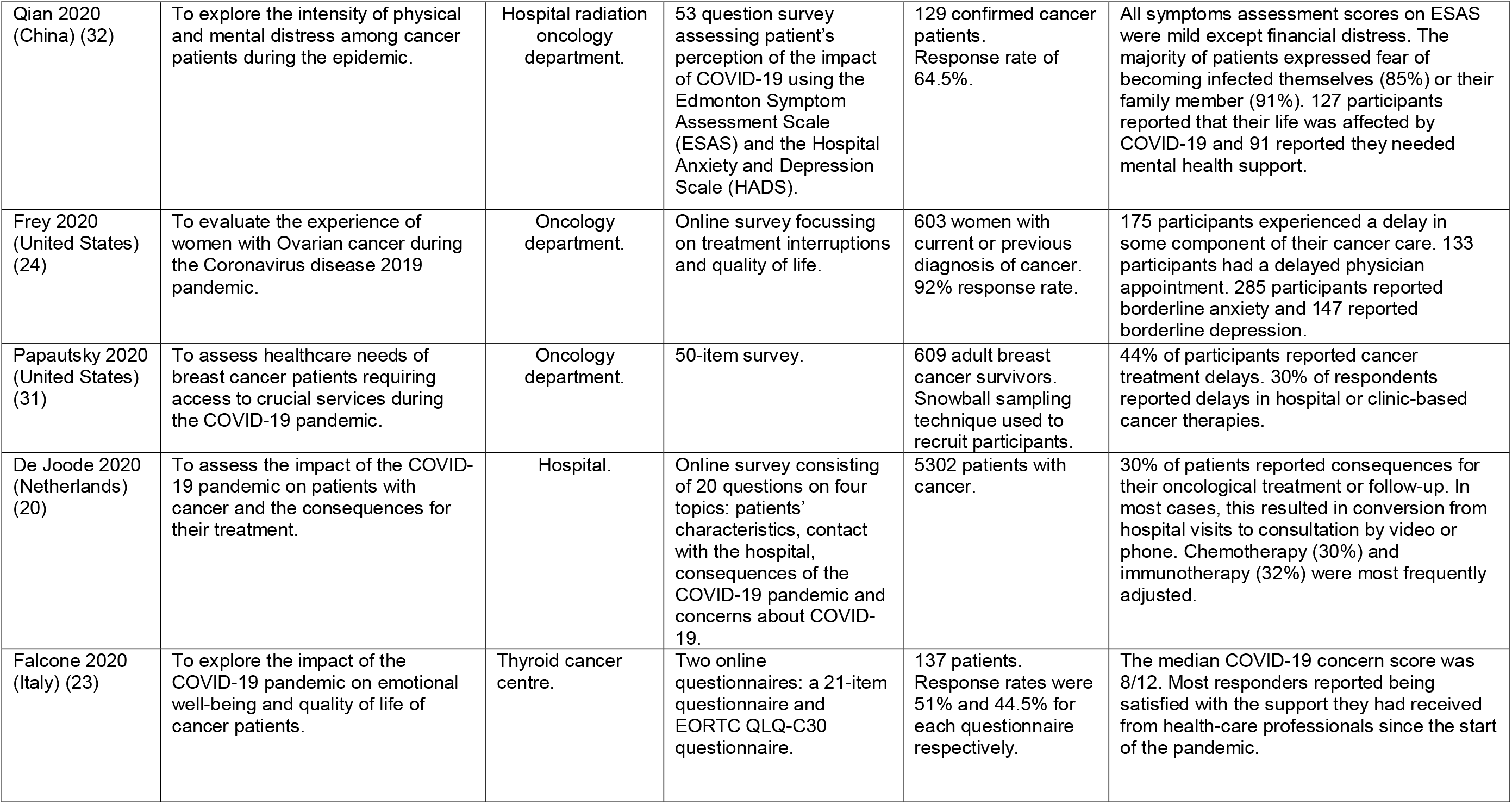
Characteristics of quantitative studies and key results.

**Figure 1:**
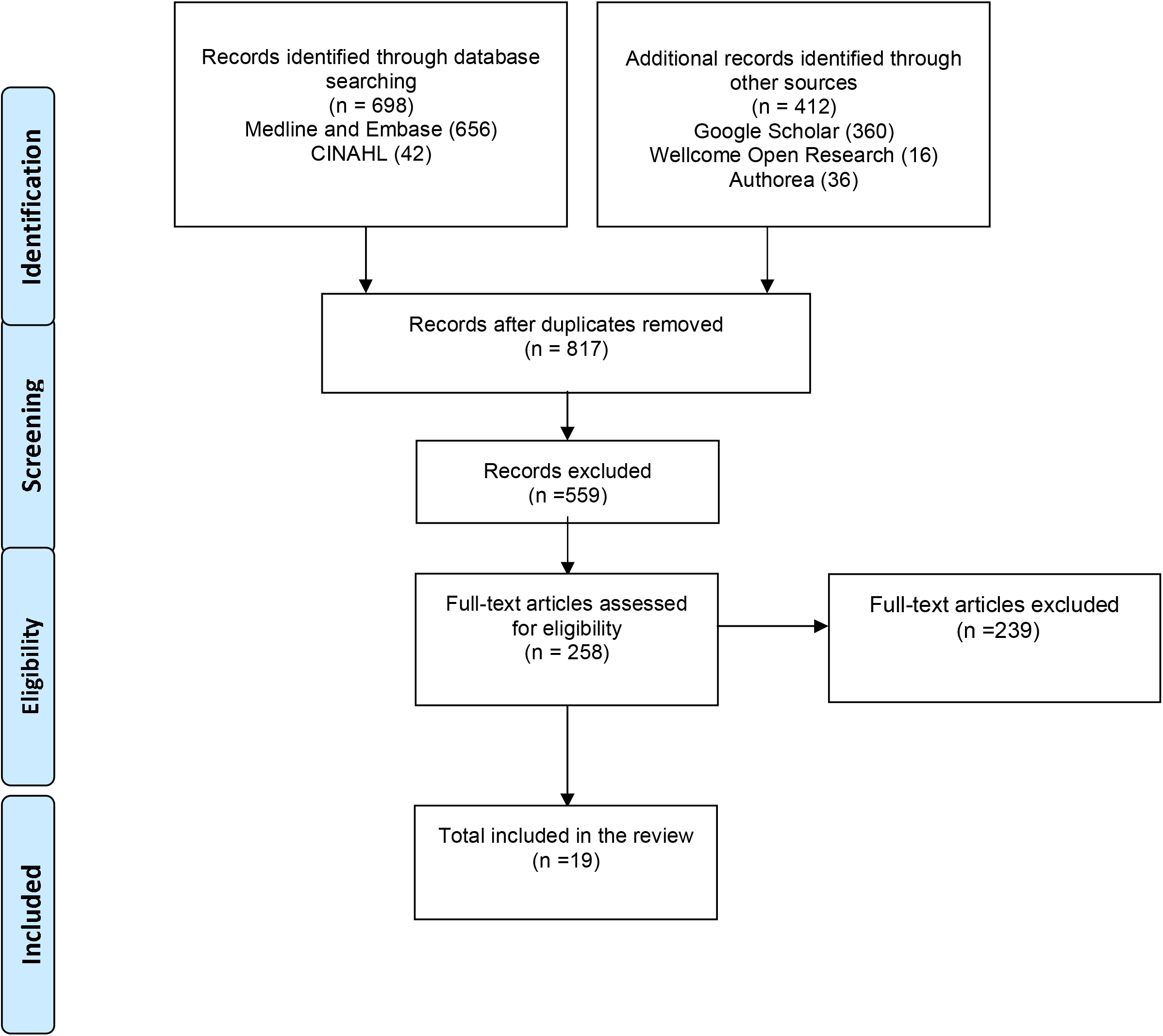
PRISMA flowchart.

### Results of thematic synthesis

Seven key categories of synthesised data and several themes were identified across the nineteen included studies. These related to-experiences of accessing cancer screening and diagnosis; experiences of accessing cancer treatment and care services; communication in relation to healthcare; perceived risks of infection; anxiety and fear; adverse impact on personal life, family and finances; caregivers specific concerns; and resilience and coping mechanisms (table 3).

**Table 3:** Emerging categories and themes from thematic synthesis.

| **Categories** | **Themes** |
| --- | --- |
| **I. Experiences of accessing cancer screening and diagnosis.** | • Oncology appointment cancellations.<br>• Reductions in referrals.<br>• Screening non-attendance.<br>• Delays in diagnosis. |
| **II. Experiences of accessing cancer treatment and care services.** | • Postponement/delays to treatment schedules.<br>• Medicine shortages.<br>• Not allowing visitors in hospitals.<br>• Changes to the prioritisation of care.<br>• Cancellations of psychological counselling sessions.<br>• Changes to nursing practices. |
| **III. Communication in relation to cancer care.** | • Views and experiences of remote consultations. |
| **IV. Perceived risks of infection, anxiety and fear.** | • Fear of self and family members contracting the virus.<br>• Fear of incomplete cancer treatment.<br>• Concerns about contracting COVID-19 and the associated consequences.<br>• Depression and changes in cognitive and emotional functioning.<br>• Government shielding letter. |
| **V. Adverse impact on personal life, family and finances.** | • Consequences of quarantine.<br>• Feelings of loneliness and isolation.<br>• Disruption to work-life.<br>• Fulfilment of end of life wishes. |
| **VI. Caregivers specific concerns.** | • Stressful aspects of care giving during a pandemic.<br>• Grieving the loss of a relative.<br>• Caregivers own health concerns. |
| **VII. Resilience and coping mechanisms** | • Feelings of peace and belonging.<br>• Greater appreciation of life.<br>• Time to develop resilient coping mechanisms. |

#### (I) Experiences of accessing cancer screening and diagnostic services

Disruption to cancer screening and diagnosis was a common theme reported in the included studies. A cross-sectional survey assessing the experiences of sarcoma patients at two of the largest specialist sarcoma centres in Europe reported that one-third of patients experienced postponement of appointments or scans by at least three months [35]. Patients also reported cancellations of routine follow-up clinic appointments in two studies, each conducted in the UK and US [31, 34]. Patients expressed feelings of anxiety and fear due to postponement of cancer-related laboratory tests and diagnostic imaging in two studies [24, 31]. This overwhelming sense of anxiety amongst patients was demonstrated further in one study where patients described the COVID-19 pandemic as overshadowing the needs of families and severely affecting the quality and scope of cancer services. Some patients described their experience of COVID-19 prevention taking precedence over cancer treatment and care [29].

#### (II) Experiences of accessing cancer treatment and care services

Participants in the majority of the included studies reported experiences of treatment delays [17,18,18,20,22,24,25-31]. Reasons ranged from hospital cancellations, city lockdowns and swabbing requirements. Deferral of radiotherapy dates and long waiting hours beyond scheduled appointment times was a problem cited by a number of participants in one study conducted in India [30]. Another study conducted in Italy reported that radiation, infusion therapies and surgical tumour removal experienced the most disruption [31]. The impact of the pandemic on cancer surgery was demonstrated in one study conducted in the US, where 10 of the included 38 participants scheduled for surgical treatment for ovarian cancer experienced delays [24]. A study held at a tertiary-care referral hospital in Italy reported a median surgery waiting time of approximately 53 days (IQR: 35-72 days); reported to be a sharp increase on pre-pandemic waiting times [26].

Whilst the interruption of treatment was most apparent, one study based in India explored other implications of the pandemic including the deferral of advice from the nutritionist, unavailability of peer group support services and psychological counselling sessions [30]. Although some of the participants had access to online counselling services, appointments were scarce.

Emotional distress was common in patients who had experienced disruption to cancer treatment [28]. Uncertainty surrounding medication availability and the lack of nursing care available for patients who were living alone added to their worries [22, 30]. For example, patients with cervical and oral cancer struggled to source certain dressing materials and so were left with defective dressings [22]. Visitors were prohibited from entering wards or accompanying the patient during appointments. In a study conducted at a palliative care centre in the US, participants likened the experience to being ‘prisoners in a cage’ [21].

#### (III) Communication in relation to cancer care

Disruptions in regular communications between patients and health services were commonly reported including the use of remote forms of communications using video technology or telephone [17,20,22,30,35]. Patients identified positive aspects of telecommunication for example, the ease of accessing care from the privacy and comfort of one’s home and the ability to maintain physical distance [22]. The majority of patients in one study hoped for continuation of online services post-pandemic [25]. In another study, almost two-thirds of patients felt that despite virtual means, they were still able to contact the healthcare team and so expressed feelings of reassurance [35]. However in one study conducted in India, patients reported difficulties in booking a virtual appointments and unpredictable network issues [30].

#### (IV) Perceived risk of infection, anxiety and fear

Feelings of anxiety and fear surrounding the pandemic was common with eighteen studies referencing changes in emotional and psychological functioning [17-30,32-35]. Approximately 55% of the 204 participants in a study conducted in a tertiary-institution study felt unsafe to visit the hospital [17]. Furthermore, cancer patients appeared overall more afraid about the complications which may arise from contracting COVID-19 [17] Prospects of not being able to say farewell to family and friends in case of dying were also reported [33]. References were also made to fears surrounding family members contracting the virus and patients expressed worries and concerns about treatment delays due to the postponement of elective procedures [32]. More than half of the participants in a US-led study disclosed a new-onset of borderline anxiety or depression [24]. Nearly a quarter (23% of the 204) participants in one study were in receipt of the government ‘shielding’ advice [34].

Two studies conducted in Saudi Arabia and Iran examined anxiety responses amongst children and their caregivers [17,29]. Parents reported fears surrounding COVID-19 mortality rates and expressed concerns about the high transmissibility and limited knowledge surrounding the virus [29]. In one of these studies [17], over 67% of parents reported the onset of new behavioural occurrences amongst their children since the pandemic. Parents were worried about the negative effects of the pandemic on children’s mental and physical health, both now and in the long-term.

#### (V) Adverse impact on personal life, family and finances

Ten studies reported wider implications of the pandemic on cancer patient’s personal life, their family and the potential financial consequences [17, 19, 22, 24, 28, 29, 32-35]. Similarly, impact on social activities due to lockdowns were also described. Loneliness fuelled patients worries surrounding cancer and previously, social outings had offered them a healthy distraction [33]. Concerns surrounding loss of income and employment instability for family members were reported [35]. Patients in palliative care expressed frustration and fear at the possibility of not being able to fulfil their last wishes; for example, spending their last days surrounded by family, as social isolation and travel restrictions had made this difficult [22].

#### (VI) Caregiver specific concerns

Concerns raised by caregivers were reported in two studies [22, 33]. In one study conducted in the Netherlands, more than half of the participants reported that they were worried about infecting cancer patients they were caring with COVID-19 [33]. In the second study, caregivers reported feelings of guilt due to their inability to ease their relatives’ suffering and in some cases were reluctant to go near the patient and change dressings due to worries about transmitting COVID-19 [22]. Moreover, caregivers reported that once lockdown commenced, they experienced feelings of helplessness as social distancing impacted their ability to provide adequate care [22].

Employment uncertainty was often reported to divert caregivers attention from caring for their relative and for some, the financial strain of the pandemic severely impacted their ability to address the basic needs of their patients. This ranged from sorting ambulance travel to providing necessities such as nutritious food; worsening feeling of guilt [22]. There was also expression of the difficulty arising from not being able to find clear information about COVID-19 and its potential impact on already vulnerable cancer patients [22].

#### (VII) Developing resilience and coping mechanisms

Patient and/or carer strategies to developing resilience and coping mechanisms were described by five studies [19, 28, 35, 29, 33]. For example, in one paediatric hospital based study, patients reported that together with their families that they altered their attitudes of nervousness and fear in order to regain control of living with the virus. They also developed strategies to deal with the anxiety associated with COVID-19 and cancer, and adapted themselves to live accordingly [29]. In two further studies, patients emphasised their appreciation for everyday life and reported feeling content as a result of lockdown [28, 33]. The lockdown offered them the time to reflect positively and focus on the ‘silver lining’ with many reporting that the daily overload they had experienced from external stimuli prior to the pandemic, had finally migrated. This created a sense of peace which seemed to ease their worries [28]. Several patients also described how the nature of their condition meant that they often spent a lot of time indoors however, with everyone being confined to their homes, they no longer felt isolated and so this reinstated a sense of belonging [33].

## Discussion

### Discussion of key findings and implications

This is the first systematic review that has explored patients and their caregivers’ views, experiences and perceived impact of the pandemic on cancer screening and care. The review has identified major themes including barriers to accessing cancer screening and diagnosis; perceived risks of infection; anxiety and fear; adverse impact on personal life, family and finances, caregivers concerns and resilience and coping mechanisms adopted by patients and carers.

The entire landscape of cancer management has changed as a result of the COVID-19 pandemic [36]. Whilst the findings from this review reinforce this, they also provide an insight into patients and carer experiences following the disruption to cancer screening and treatment. As a result of diagnostic and treatment delays, governments and health systems are expecting a surge in the number of avoidable, cancer-related deaths [37]. Participants in the included studies of this review expressed concerns surrounding incomplete treatment, complications associated with contracting COVID-19 and changes to their psychological and emotional wellbeing. The suspension of cancer screening, cancellation of routine oncology appointments and postponement of treatment have each been associated with increased feelings of stress and anxiety; suggesting that there is a critical gap in disaster preparedness [38]. Consequently, vulnerable patients are left at greater risk of poor cancer outcomes with additional implications for mental health, symptom control and quality of life.

Perceived risk of infection, anxiety and fear when using cancer services was identified as a common theme. Such theme was prominent in studies conducted in low-and-middle income countries. For example, a study conducted in Gaza reported that prior to COVID-19, only one toilet was available for public use in the oncology department and with cancer patients expressing their fears about the lack of social distancing and, close proximity [18]. The World Health Organization (WHO) has also voiced concern about the lack of personal protective equipment (PPE), drug deficit and power supply problems [39]. The repercussions of resource availability on cancer treatment outcomes outlined above are further supported by current research which highlights the unequal impact of the Coronavirus pandemic [40] however, further in-depth review is required to explore this further.

Lessons learnt from this pandemic should become an integral part of oncology practice and thus ensure that there is the continuum of cancer care despite external challenges. Future pandemic preparedness is necessary in order to minimise the disruption experienced by both cancer patients and their caregivers. Efforts should firstly be placed on restoring cancer services with the prioritisation of screening, early detection and diagnosis according to patients COVID-19 and cancer risk. As primary care is focal to diagnosis, it is vital that patients with symptoms are encouraged, and therefore feel comfortable with seeking medical help and are assessed in a timely manner [41]. Remote consulting comes with many communication challenges for example, ‘missed cues.’ These would normally be more apparent in face-to-face consultations.

Introducing innovations such as triage tests for patients reporting a specific set of symptoms may help to address such communication challenges and ensure that prompt referrals are made [41]. The consequent delay of treatment due to COVID-related measures should be considered on an individual, case by case basis for each patient; ensuring that critical patients receive adequate therapy. Alongside this, the public health implications of potentially delaying treatment versus COVID-19 prevention should be assessed and seemingly a balance is required.

Internationally, the professional cancer societies including The American Society of Clinical Oncology (ASCO) have produced guidance in relation to safe provision and continuity of cancer care during the pandemic [42,43]. While these guidance advocate greater flexibility and alternative treatment options to suit patient circumstances, the impact on personal, psychological and financial wellbeing on patients need to be considered. Patient centred communications from health service providers to address patient fears of contracting the virus while on treatment and reluctance to use health services are essential.

Throughout the pandemic, it is necessary to evaluate detection rates, treatment uptake and outcomes so that valid pre-pandemic comparisons can be made. It is also imperative to consider healthcare professionals’ and wider stakeholders’ experiences of delivering cancer services.

### Strengths and limitations

Although we adopted rigorous and systematic approach to conducting the review, our study has some limitations. As the coronavirus pandemic is such a recent and rapidly advancing area of research, the date of coverage was very limited i.e.one year. As this is an international systematic review, data has been synthesised from a range of countries. However, healthcare systems can vary considerably and so caution should be exercised when considering the generalisability of the findings.

## Conclusion

This systematic review suggests that globally, postponement and delays in the cancer screening and treatment, drug shortages and inadequate nursing care were commonly experienced these contributed to anxiety and fear amongst patients and carers. Hospital closures, lockdowns and patient reluctance to use services contributed to the treatment delays and cancellations. Patients had to undergo treatment or experience cancer journey on their own due to infection measures in treatment centres and shielding at home. Caregivers were reported

to be anxious about infecting cancer patients and financial concerns due to COVID-19 affected their ability to care. There is a need for sustained effort in continuation of essential cancer care services during the time of the pandemic. Clear communication from cancer services alongside activation of patient outreach services is imperative. Future research and service models should incorporate patient and caregiver views identified from this study.

## Supporting information

Electronic supplement

## Data Availability

The data underlying this article are available in the article and supplementary materials submitted with the manuscript.

## Declarations

### Ethics approval and consent to participate

Not applicable

### Consent for publication

Not applicable

## Authors’ contributions

VP was the principal investigator of the study and SD was the lead researcher. VP and DS originated the study idea. VP, MAH and SD conducted the literature search. EC and DS provided expert input into quality assessment, data extraction and analysis. SD undertook data extraction and quality assessment, VP undertook duplicate, independent assessment and quality checks.

SD produced the first draft of the manuscript to which all authors contributed with expert input and comments. All authors agree to the final version of the manuscript.

## Acknowledgements

We would like to thank the University of Birmingham Library services for their support in the search process.

## Abbreviations

ASCO: The American Society of Clinical Oncology
CASP: Critical Appraisal Skills Programme
NIH: National Institute of Health
PPE: Personal Protective Equipment
PRISMA: Preferred Reporting Items for Systematic Review and Meta-analysis

