## Supplementary material for "Cancer services during the COVID-19 pandemic: systematic review of patients’ and caregivers’ experiences": Electronic supplement

**Supplementary material 1**

**Search strategy: Ovid Medline & Embase**

1	cancer.mp. or exp *Neoplasms/ (7938965)

2	Medical Oncology/ or cancer care.mp. (164669)

3	exp *"Early Detection of Cancer"/ (16632)

4	cancer screening.mp. (120350)

5	cancer treatment.mp. (114993)

6	cancer chemotherapy.mp. (281128)

7	Coronavirus Infections/ (31794)

8	(COVID-19 or COVID19).mp. (79504)

9	coronavirus pandemic.mp. (1003)

10	access.mp. or Access to Information/ (776246)

11	Health Services Accessibility/ or service access.mp. (126429)

12	perspective$.mp. or "Attitude of Health Personnel"/ (841294)

13	experience$.mp. (2483723)

14	Attitude/ or attitude.mp. (754140)

15	view$.mp. (1003904)

16	1 or 2 or 3 or 4 or 5 or 6 (7965487)

17	7 or 8 or 9 (84883)

18	10 or 11 or 12 or 13 or 14 or 15 (5119959)

19	16 and 17 and 18 (909)

20	remove duplicates from 19 (712)

21	limit 20 to english language (698)

22	limit 21 to yr="2020 -Current" (695)

**Supplementary material 2**

**PRISMA check-list^1^.**

| **Section/topic** | **#** | **Checklist item** | **Reported in section** |
| --- | --- | --- | --- |
| **TITLE** | | |  |
| Title | 1 | Identify the report as a systematic review, meta-analysis, or both. | Title |
| **ABSTRACT** | | |  |
| Structured summary | 2 | Provide a structured summary including, as applicable: background; objectives; data sources; study eligibility criteria, participants, and interventions; study appraisal and synthesis methods; results; limitations; conclusions and implications of key findings; systematic review registration number. | Abstract |
| **INTRODUCTION** | | |  |
| Rationale | 3 | Describe the rationale for the review in the context of what is already known. | Introduction |
| Objectives | 4 | Provide an explicit statement of questions being addressed with reference to participants, interventions, comparisons, outcomes, and study design (PICOS). | Introduction |
| **METHODS** | | |  |
| Protocol and registration | 5 | Indicate if a review protocol exists, if and where it can be accessed (e.g., Web address), and, if available, provide registration information including registration number. | Abstract/introduction |
| Eligibility criteria | 6 | Specify study characteristics (e.g., PICOS, length of follow-up) and report characteristics (e.g., years considered, language, publication status) used as criteria for eligibility, giving rationale. | Methods |
| Information sources | 7 | Describe all information sources (e.g., databases with dates of coverage, contact with study authors to identify additional studies) in the search and date last searched. | Methods |
| Search | 8 | Present full electronic search strategy for at least one database, including any limits used, such that it could be repeated. | Methods/electronic supplementary material 1/figure 1 |
| Study selection | 9 | State the process for selecting studies (i.e., screening, eligibility, included in systematic review, and, if applicable, included in the meta-analysis). | Method |
| Data collection process | 10 | Describe method of data extraction from reports (e.g., piloted forms, independently, in duplicate) and any processes for obtaining and confirming data from investigators. | Method |
| Data items | 11 | List and define all variables for which data were sought (e.g., PICOS, funding sources) and any assumptions and simplifications made. | Method |
| Risk of bias in individual studies | 12 | Describe methods used for assessing risk of bias of individual studies (including specification of whether this was done at the study or outcome level), and how this information is to be used in any data synthesis. | Method, electronic supplementary materials 3 and 4 |
| Summary measures | 13 | State the principal summary measures (e.g., risk ratio, difference in means). | N/A |
| Synthesis of results | 14 | Describe the methods of handling data and combining results of studies, if done, including measures of consistency (e.g., I^2^) for each meta-analysis. | Metods |

| **Section/topic** | **#** | **Checklist item** | **Reported on page #** |
| --- | --- | --- | --- |
| Risk of bias across studies | 15 | Specify any assessment of risk of bias that may affect the cumulative evidence (e.g., publication bias, selective reporting within studies). | Electronic supplementary materials 3 and 4 |
| Additional analyses | 16 | Describe methods of additional analyses (e.g., sensitivity or subgroup analyses, meta-regression), if done, indicating which were pre-specified. | N/A |
| **RESULTS** | | |  |
| Study selection | 17 | Give numbers of studies screened, assessed for eligibility, and included in the review, with reasons for exclusions at each stage, ideally with a flow diagram. | Results/ figure 1 |
| Study characteristics | 18 | For each study, present characteristics for which data were extracted (e.g., study size, PICOS, follow-up period) and provide the citations. | Tables 1 and 2 |
| Risk of bias within studies | 19 | Present data on risk of bias of each study and, if available, any outcome level assessment (see item 12). | Electronic supplementary materials 3 and 4 |
| Results of individual studies | 20 | For all outcomes considered (benefits or harms), present, for each study: (a) simple summary data for each intervention group (b) effect estimates and confidence intervals, ideally with a forest plot. | N/A |
| Synthesis of results | 21 | Present results of each meta-analysis done, including confidence intervals and measures of consistency. | Table 3 |
| Risk of bias across studies | 22 | Present results of any assessment of risk of bias across studies (see Item 15). | Electronic supplementary materials 3 and 4 |
| Additional analysis | 23 | Give results of additional analyses, if done (e.g., sensitivity or subgroup analyses, meta-regression [see Item 16]). | N/A |
| **DISCUSSION** | | |  |
| Summary of evidence | 24 | Summarize the main findings including the strength of evidence for each main outcome; consider their relevance to key groups (e.g., healthcare providers, users, and policy makers). | Discussion |
| Limitations | 25 | Discuss limitations at study and outcome level (e.g., risk of bias), and at review-level (e.g., incomplete retrieval of identified research, reporting bias). | Discussion |
| Conclusions | 26 | Provide a general interpretation of the results in the context of other evidence, and implications for future research. | Discussion |
| **FUNDING** | | |  |
| Funding | 27 | Describe sources of funding for the systematic review and other support (e.g., supply of data); role of funders for the systematic review. | Manuscript data |

^1^Moher D, Liberati A, Tetzlaff J, Altman DG, The PRISMA Group (2009). Preferred Reporting Items for Systematic Reviews and Meta-Analyses: The PRISMA Statement. PLoS Med 6(7): e1000097. doi:10.1371/journal.pmed1000097

N/A= Not applicable.

**Electronic supplementary material 3: Quality assessment of Qualitative studies using CASP.**

|  | **Study ID** | | | | |
| --- | --- | --- | --- | --- | --- |
|  | Dhavale 2020 (22) | Mirlashari 2020 (29) | Hyland 2020 (28) | AlWaheidi 2020 (18) | Casanova 2020 (19) |
| **Quality assessment criteria** |  |  |  |  |  |
| Was there a clear statement of the aims of the research? | Yes | Yes | Yes | No | Yes |
| Is a qualitative methodology appropriate? | Yes | Yes | Yes | Can’t tell | Can’t tell |
| Was the research design appropriate to address the aims of the research? | Yes | Yes | Yes | Can’t tell | Yes |
| Was the recruitment strategy appropriate to the aims of the research? | Can’t tell | No | No | Can’t tell | Can’t tell |
| Was the data collected in a way that addressed the research issue? | Can’t tell | Yes | Yes | Can’t tell | Can’t tell |
| Has the relationship between researcher and participants been adequately considered? | No | Yes | Yes | Can’t tell | No |
| Have ethical issues been taken into consideration? | Can't tell | Can't tell | Can't tell | Can't tell | Can't tell |
| Was the data analysis sufficiently rigorous? | No | Yes | Can’t tell | Can’t tell | Yes |
| Is there a clear statement of findings? | Yes | Yes | Yes | Yes | Yes |

**Electronic supplementary material 4: Quality assessment of Quantitative studies**

|  | AlShahrani 2020 (17) | Swainston 2020 (34) | Desideri 2020 (21) | Guven 2020 (27) | Schellekens 2020 (33) | Greco 2020 (26) | Mitra 2020 (30) | Younger 2020 (35) | Ghosh 2020 (25) | Qian 2020 (32) | Frey 2020 (24) | Papautsky (31) | De Joode 2020 (20) | Falcone 2020 (23) |
| --- | --- | --- | --- | --- | --- | --- | --- | --- | --- | --- | --- | --- | --- | --- |
| **Quality assessment criteria** |  |  |  |  |  |  |  |  |  |  |  |  |  |  |
| Was the research question or objective in this paper clearly stated? | Yes | Yes | Yes | Yes | Yes | Yes | Yes | Yes | Yes | Yes | Yes | Yes | Yes | Yes |
| Was the study population clearly specified and defined? | Yes | Yes | Yes | Yes | Yes | Yes | Yes | Yes | Yes | Yes | Yes | Yes | Yes | Yes |
| Was the participation rate of eligible persons at least 50%? | NR | NR | NR | Yes | No | Yes | No | No | NR | Yes | Yes | NR | NR | No |
| Were all the subjects selected or recruited from the same or similar populations (including the same time period)? | Yes | Yes | Yes | Yes | Yes | Yes | Yes | Yes | Yes | Yes | Yes | Yes | NR | Yes |
| Were inclusion and exclusion criteria for being in the study prespecified and applied uniformly to all participants? | Yes | Yes | Yes | No | No | Yes | No | No | Yes | No | No | Yes | No | No |
| Was a sample size justification, power description, or variance and effect estimates provided? | No | No | No | No | No | No | No | No | No | No | No | No | No | No |

NR= Not reported
